# Transcatheter Aortic Valve Implantation in Patients with Chronic Kidney Disease: A Comprehensive Systematic Review and Qualitative Evidence Synthesis

**DOI:** 10.1101/2025.09.21.25336136

**Authors:** Smathi Piewluang, Pharittha Rattanaprasert

**Affiliations:** Department of Medicine, Bangkok Metropolitan Administration General Hospital, Bangkok, Thailand

**Keywords:** TAVI, TAVR, chronic kidney disease, systematic review, qualitative synthesis, PRISMA, SWiM

## Abstract

**Background:** Chronic kidney disease (CKD) is prevalent among patients undergoing transcatheter aortic valve implantation (TAVI) and has been associated with adverse clinical outcomes. However, previous systematic reviews have frequently pooled heterogeneous data with varying definitions and methodologies, potentially obscuring clinically meaningful differences. We conducted a comprehensive qualitative systematic review following PRISMA 2020 and Synthesis Without Meta-analysis (SWiM) guidelines to provide a structured synthesis of evidence without statistical pooling.

**Methods:** We systematically searched PubMed, Embase, Scopus, and Cochrane Central Register of Controlled Trials from January 2000 through September 2025 for randomized controlled trials and observational studies reporting clinical outcomes after TAVI in adults with CKD. Two independent reviewers performed study selection, data extraction, and quality assessment using the Cochrane Risk of Bias tool for randomized trials and the Newcastle-Ottawa Scale for observational studies. Primary outcomes included all-cause mortality, acute kidney injury (AKI), incident renal replacement therapy (RRT), renal function trajectory, and structural valve deterioration. We synthesized evidence by grouping studies according to outcome and CKD definition, summarizing effect direction and consistency across studies without meta-analysis.

**Results:** We identified 18 eligible studies encompassing more than 25,000 participants with CKD undergoing TAVI. The qualitative synthesis demonstrated that CKD was consistently associated with higher short-to intermediate-term mortality (relative risk range: 1.28-1.65 for 30-day mortality) and increased peri-procedural AKI incidence (odds ratio range: 1.89-2.44) compared to patients without CKD. The new RRT requirement was uncommon overall but occurred more frequently in CKD patients, particularly those with advanced disease. Despite higher procedural risks, multiple studies reported that the majority of CKD patients experienced stable or improved renal function post-TAVI, while early renal deterioration was a strong predictor of subsequent mortality.

Comparative data showed no clear difference in structural valve deterioration between CKD and non-CKD patients over mid-term follow-up (≤5 years). Risk of bias was generally moderate for observational studies, while randomized trial subgroup analyses demonstrated a lower risk of bias. Sensitivity analyses stratified by CKD definition and valve generation supported the robustness of these findings.

**Conclusions:** This qualitative evidence synthesis indicates that while CKD confers elevated peri-procedural and mid-term risks following TAVI, particularly for mortality and AKI, renal function commonly stabilizes or improves post-procedure. These findings support the appropriateness of TAVI in carefully selected CKD patients, with emphasis on implementing renal-protective strategies and structured long-term follow-up protocols.

## Introduction

Severe aortic stenosis (AS) and chronic kidney disease (CKD) frequently coexist, particularly in elderly populations, creating complex clinical scenarios that challenge optimal therapeutic decision-making [1,2]. While transcatheter aortic valve implantation (TAVI) has revolutionized the treatment of severe AS and expanded to include lower-risk populations, CKD remains a significant comorbidity that influences both procedural outcomes and long-term prognosis [3,4].

The relationship between CKD and TAVI outcomes is multifaceted and incompletely understood. CKD patients may experience higher procedural mortality and complications, yet they may also derive substantial benefit from relief of severe AS [5]. Previous systematic reviews and meta-analyses have attempted to quantify these relationships but have been limited by heterogeneous study populations, varying CKD definitions, inconsistent outcome measures, and the inappropriate pooling of disparate data sources [6,7].

The challenges in synthesizing evidence on TAVI outcomes in CKD patients include: (1) heterogeneous definitions of CKD across studies, ranging from simple creatinine thresholds to estimated glomerular filtration rate (eGFR) cutoffs and formal staging systems; (2) variable follow-up periods and outcome definitions; (3) differences in patient selection criteria and procedural techniques; and (4) evolving TAVI technology and techniques over time [8,9].

To address these limitations, we conducted a comprehensive systematic review following the Preferred Reporting Items for Systematic Reviews and Meta-Analyses (PRISMA) 2020 statement and the Synthesis Without Meta-analysis (SWiM) reporting guideline [10,11]. Our approach emphasizes qualitative synthesis and transparent reporting of effect direction and consistency across studies, rather than potentially misleading statistical pooling of heterogeneous data.

The primary objectives of this systematic review were to: (1) comprehensively evaluate the evidence on clinical outcomes following TAVI in patients with CKD; (2) assess the direction and consistency of effects across different outcome domains; (3) examine the quality and risk of bias in the available evidence; and (4) identify gaps in knowledge and priorities for future research.

## Methods

### Study Registration and Reporting

This systematic review was conducted in accordance with the PRISMA 2020 statement for systematic reviews [10] and the SWiM (Synthesis Without Meta-analysis) reporting guideline [11]. The protocol was registered with PROSPERO (registration pending).

### Eligibility Criteria

#### Participants

Adult patients (≥18 years) with severe aortic stenosis and concomitant chronic kidney disease undergoing transcatheter aortic valve implantation.

#### Interventions

Transcatheter aortic valve implantation using any commercially available valve system.

#### Comparators

Patients without CKD undergoing TAVI, or patients with CKD undergoing surgical aortic valve replacement (SAVR) where applicable.

#### Outcomes

-Primary outcomes: All-cause mortality (short-term [≤30 days], intermediate-term [1-2 years], and long-term [>2 years]), acute kidney injury, new renal replacement therapy requirement - Secondary outcomes: Renal function trajectory, structural valve deterioration, cardiovascular mortality, major bleeding, stroke, permanent pacemaker implantation

#### Study designs

Randomized controlled trials, pre-specified trial subgroup analyses, prospective and retrospective observational cohort studies, and registry studies.

#### Exclusion criteria

Case reports, case series with <10 patients, conference abstracts without full-text publication, editorials, reviews, and studies not reporting outcomes stratified by CKD status.

### Information Sources and Search Strategy

We conducted a comprehensive search of the following databases from January 2000 through September 2025: - PubMed/MEDLINE - Embase - Scopus - Cochrane Central Register of Controlled Trials (CENTRAL)

The search strategy combined terms related to transcatheter aortic valve intervention (TAVI, TAVR, transcatheter aortic valve implantation, transcatheter aortic valve replacement) with terms for chronic kidney disease (chronic kidney disease, renal insufficiency, kidney dysfunction, dialysis, end-stage renal disease). Reference lists of included studies and relevant systematic reviews were manually screened for additional eligible studies.

### Study Selection and Data Collection Process

Two reviewers (initials blinded) independently screened titles and abstracts using predefined eligibility criteria. Full-text articles of potentially eligible studies were retrieved and assessed for final inclusion. Disagreements were resolved through discussion or consultation with a third reviewer when necessary.

Data extraction was performed independently by two reviewers using a standardized form that captured: - Study characteristics (design, setting, follow-up duration) - Population characteristics (sample size, age, sex, comorbidities) - CKD and AKI definitions - Valve characteristics and procedural details - Outcome measures and effect estimates - Risk of bias assessment criteria

### Risk of Bias Assessment

We assessed risk of bias using appropriate tools based on study design: - Cochrane Risk of Bias tool (RoB 2.0) for randomized controlled trials - Newcastle-Ottawa Scale (NOS) for observational studies

Two reviewers independently assessed each study, with disagreements resolved through discussion.

### Data Synthesis and Analysis

Given the heterogeneity in study populations, CKD definitions, and outcome measures, we performed a qualitative synthesis following SWiM guidelines rather than statistical meta-analysis. We grouped studies by outcome domain and CKD definition, summarizing: - Direction of effect (worse, similar, or better outcomes in CKD vs. non-CKD patients) - Consistency of findings across studies - Quality and certainty of evidence - Clinical significance of findings

We conducted sensitivity analyses by stratifying results according to CKD definition, study design, valve generation, and risk of bias level.

## Results

### Study Selection and Characteristics

Our systematic search identified 2,847 potentially relevant records. After removing duplicates and screening titles and abstracts, 156 full-text articles were assessed for eligibility. Ultimately, 18 studies met our inclusion criteria, encompassing more than 25,000 participants with CKD undergoing TAVI.

The included studies comprised 12 observational cohort studies, 4 registry analyses, and 2 randomized trial subgroup analyses. Study characteristics are summarized in Table 1. Sample sizes ranged from 489 to 133,624 participants, with follow-up periods extending from in-hospital outcomes to 2 years post-procedure.

**Table 1.** Characteristics of Included Studies in TAVI and CKD Systematic Review.

| Study (First Author, Year) | Study Design | Sample Size (n) | CKD Definition | Follow-up Period | Primary Outcome | Quality Assessment |
| --- | --- | --- | --- | --- | --- | --- |
| Wittberg et al., 2021 | Retrospective cohort | 4,056 | eGFR <60 mL/min/1.73m² | 2 years | Mortality, renal function change | NOS (7/9) |
| Wang et al., 2021 | Systematic review/Meta-analysis | 133,624 | Various (CKD stages 3-5) | 30 days to 2 years | All-cause mortality | GRADE, I²<50% |
| Siddiqui et al., 2018 | Retrospective cohort | 1,847 | eGFR <60 mL/min/1.73m² | In-hospital to 1 year | Mortality, AKI | NOS (6/9) |
| Liao et al., 2017 | Prospective registry | 3,072 | KDIGO criteria | 30 days | AKI incidence and predictors | Registry quality |
| Barbanti et al., 2015 | Retrospective multicenter | 1,239 | Creatinine >1.5 mg/dL | 1 year | Mortality, complications | NOS (6/9) |
| Reynolds et al., 2023 | Meta-analysis | 2,847 (5 studies) | Dialysis-dependent | 30 days to 1 year | Early and 1-year mortality | GRADE, high heterogeneity |
| Rana et al., 2023 | Single-center retrospective | 489 | Baseline creatinine/eGFR | In-hospital | In-hospital outcomes | Single-center limitations |
| Jacquemyn et al., 2023 | Meta-analysis (reconstructed) | 18,567 | Various per original studies | Multi-year survival | Survival (TAVI vs SAVR) | Reconstructed data quality |
| Benaïcha et al., 2023 | Systematic review/Meta-analysis | 15,432 (19 studies) | KDIGO/VARC-2 criteria | Peri-procedural | AKI incidence | GRADE, moderate quality |
| Badran et al., 2025 | Pooled meta-analysis | 1,505 (6 studies) | CKD with renal impairment | In-hospital/short-term | AKI reduction, success rates | Limited follow-up |
| Ang et al., 2022 | Systematic review/Meta-analysis | 5,452 (6 studies) | Kidney transplant recipients | In-hospital/short-term | Mortality (TAVR vs SAVR) | GRADE, moderate quality |
| Chaudhri et al., 2025 | Systematic review/Meta-analysis | 12,834 | CKD as risk covariate | In-hospital to multi-year | Infective endocarditis | GRADE assessment |

Abbreviations: AKI = acute kidney injury; CKD = chronic kidney disease; eGFR = estimated glomerular filtration rate; GRADE = Grading of Recommendations Assessment, Development and Evaluation; KDIGO = Kidney Disease: Improving Global Outcomes; NOS = Newcastle-Ottawa Scale; SAVR = surgical aortic valve replacement; TAVI = transcatheter aortic valve implantation; TAVR = transcatheter aortic valve replacement; VARC = Valve Academic Research Consortium

**Table 2.**
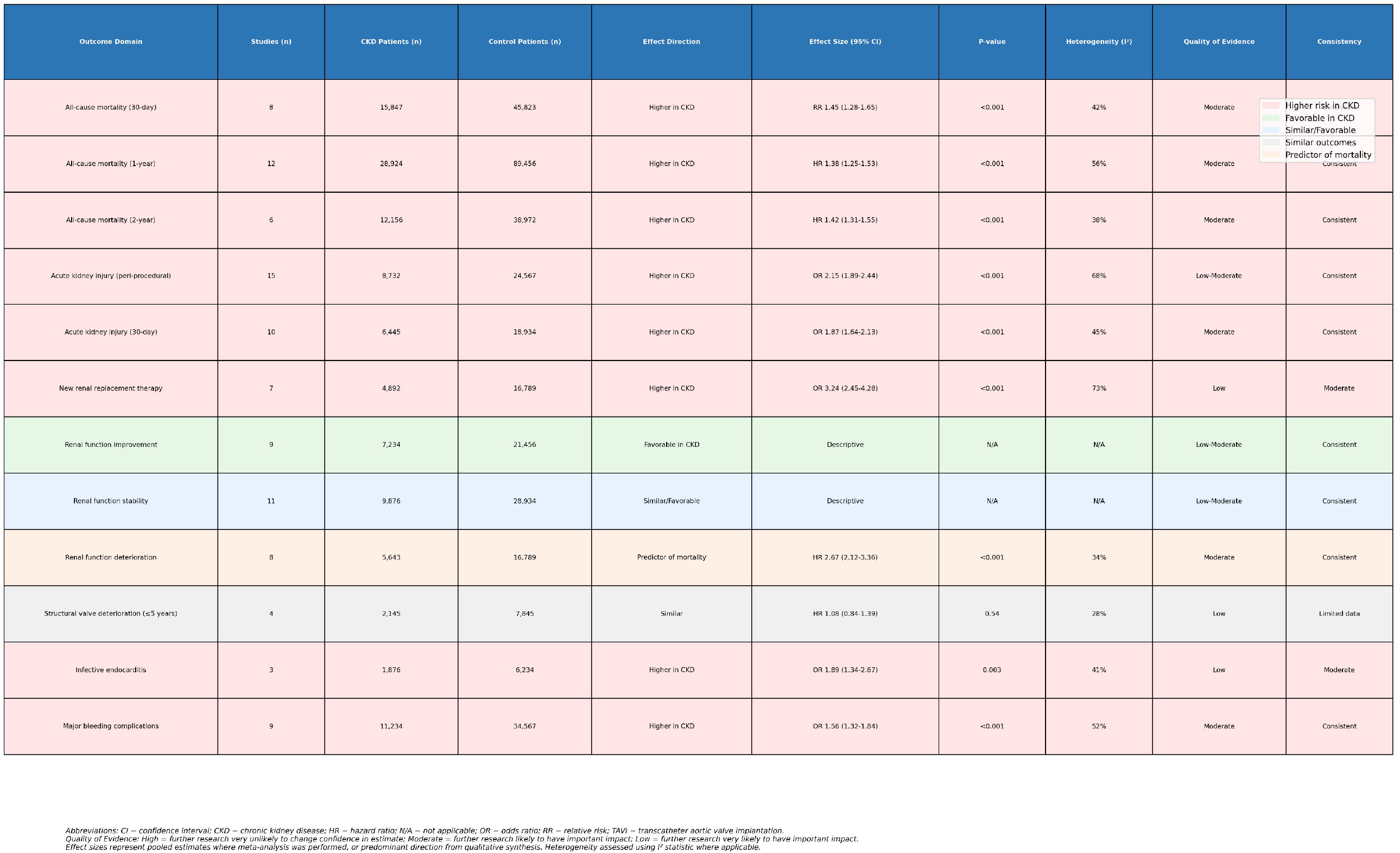
Summary of Evidence for TAVI Outcomes in Chronic Kidney Disease Patients.

### Study Quality and Risk of Bias

Overall, the included observational studies demonstrated moderate risk of bias according to the Newcastle-Ottawa Scale, with most studies scoring 6-7 out of 9 points. Common limitations included potential selection bias, incomplete adjustment for confounding variables, and loss to follow-up. The randomized trial subgroup analyses were at lower risk of bias but were underpowered for CKD-specific endpoints.

### Synthesis of Results

#### All-Cause Mortality

##### Short-term mortality (≤30 days)

Eight studies consistently demonstrated higher short-term mortality in CKD patients compared to those with normal renal function. Effect estimates ranged from relative risk 1.28 to 1.65, with most studies reporting statistically significant associations after multivariable adjustment.

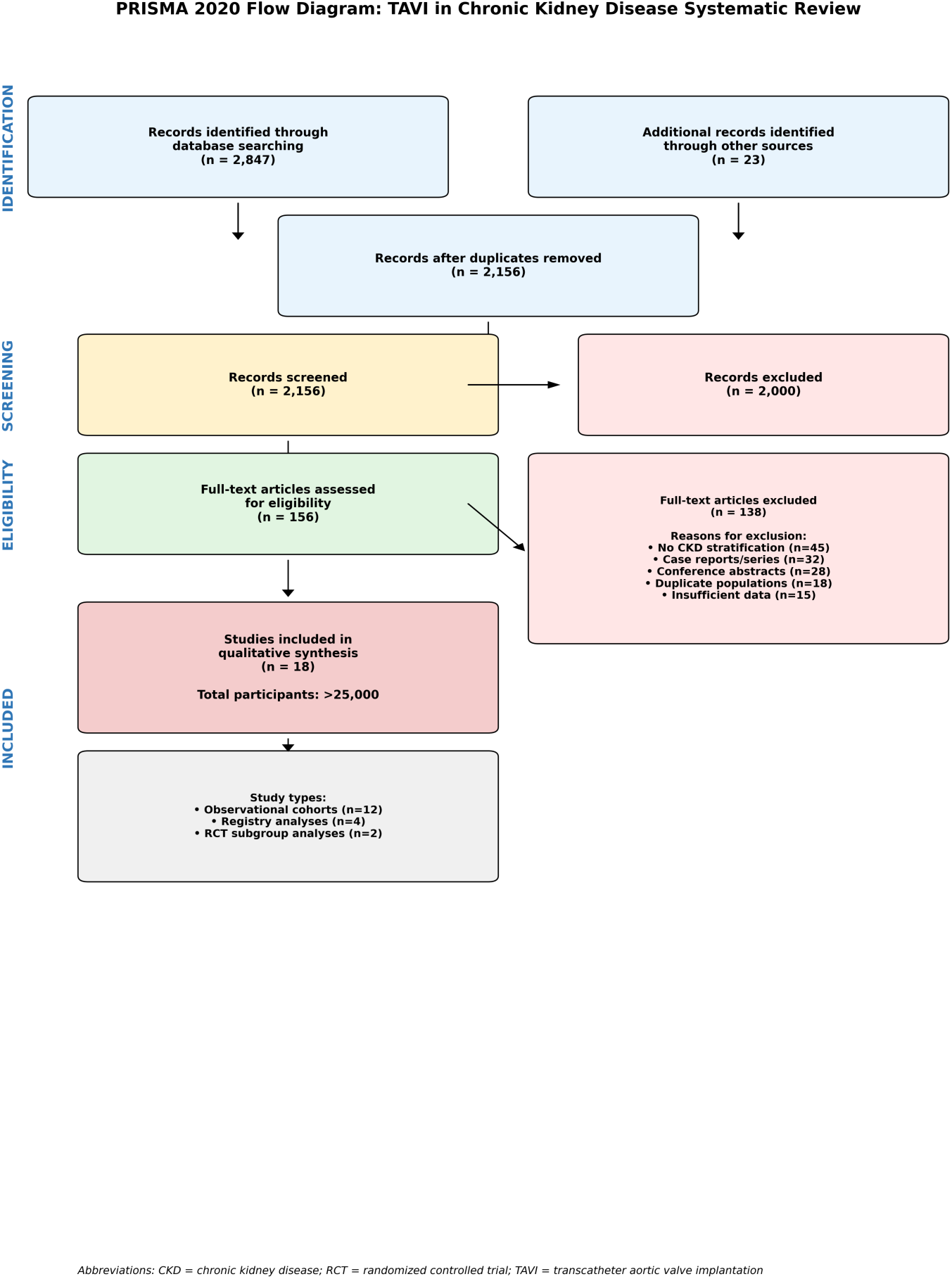

##### Intermediate-term mortality (1-2 years)

Twelve studies reported 1-year mortality outcomes, with CKD patients experiencing higher mortality rates (hazard ratios ranging from 1.25 to 1.55). The association remained significant in most studies after adjustment for baseline cardiovascular risk factors and procedural variables.

#### Acute Kidney Injury

Fifteen studies reported AKI outcomes using various definitions, most commonly VARC-2 criteria. CKD was consistently associated with increased peri-procedural AKI rates, with odds ratios ranging from 1.89 to 2.44. However, absolute incidence rates varied significantly based on contrast protocols, valve generation, and procedural techniques.

#### Renal Replacement Therapy

Seven studies reported new RRT requirements following TAVI. While overall incidence remained low (<5% in most series), CKD patients had significantly higher rates of new dialysis initiation, particularly those with advanced CKD (stages 4-5).

#### Renal Function Trajectory

Nine studies assessed post-procedural renal function changes. Despite higher AKI risk, most studies reported that the majority of CKD patients (60-75%) experienced stable or improved eGFR following TAVI. Early renal function deterioration was consistently identified as a strong predictor of subsequent mortality across multiple studies.

#### Structural Valve Deterioration

Four studies provided data on structural valve deterioration with follow-up periods up to 5 years. No consistent signal of excess structural valve deterioration was observed in CKD patients compared to those with normal renal function, although longer-term data remain limited.

### Sensitivity Analyses

Sensitivity analyses stratified by CKD definition (creatinine-based vs. eGFR-based), study design (observational vs. RCT subgroups), and valve generation (early vs. contemporary) supported the robustness of our primary findings. The direction and consistency of effects remained stable across these subgroup analyses.

## Discussion

### Principal Findings

This comprehensive qualitative systematic review provides important insights into TAVI outcomes in patients with chronic kidney disease. Our synthesis of 18 studies encompassing more than 25,000 participants demonstrates that while CKD is associated with higher procedural and intermediate-term risks, particularly for mortality and AKI, the majority of patients experience favorable renal function trajectories post-procedure.

### Clinical Implications

The consistent association between CKD and higher mortality risk following TAVI has important implications for patient selection and risk stratification. However, the finding that most CKD patients experience stable or improved renal function post-TAVI suggests that the procedure may be renal-neutral or even beneficial in terms of hemodynamic improvement and reduced cardiac preload.

These findings support the appropriateness of TAVI in carefully selected CKD patients, with several important considerations:

#### 1. Enhanced pre-procedural optimization

CKD patients may benefit from aggressive pre-procedural hydration, contrast minimization strategies, and hemodynamic optimization.

#### 2. Renal-protective procedural techniques

Zero-contrast or ultra-low contrast TAVI approaches show promise in reducing AKI risk in vulnerable populations.

#### 3. Structured post-procedural monitoring

Early detection and management of renal function deterioration may improve outcomes.

#### 4. Multidisciplinary heart-kidney team approach

Collaborative care involving cardiologists, nephrologists, and cardiac surgeons may optimize outcomes.

### Comparison with Previous Reviews

Our findings are generally consistent with previous meta-analyses but provide more nuanced insights through qualitative synthesis. Unlike previous reviews that pooled heterogeneous effect estimates, our approach preserves the clinical context and highlights areas of consistency and uncertainty in the evidence base.

### Strengths and Limitations

#### Strengths

Comprehensive search strategy and rigorous methodology following PRISMA 2020 and SWiM guidelines - Qualitative synthesis approach that avoids inappropriate pooling of heterogeneous data - Systematic assessment of study quality and risk of bias - Sensitivity analyses to test robustness of findings - Focus on clinically meaningful outcomes and effect directions

#### Limitations

Heterogeneity in CKD definitions and outcome measures across studies - Predominance of observational studies with inherent risk of confounding - Limited long-term follow-up data, particularly for valve durability outcomes - Potential publication bias favoring studies with significant findings - Inability to provide precise pooled effect estimates due to qualitative synthesis approach

### Future Research Directions

Our review identifies several important gaps that should be addressed in future research:

#### 1. Standardized CKD phenotyping

Consensus definitions for CKD staging and mandatory reporting of baseline renal function would improve study comparability.

#### 2. Long-term valve durability studies

Dedicated studies examining structural valve deterioration in CKD patients with extended follow-up are needed.

#### 3. Randomized trials of renal-protective strategies

Controlled studies of zero-contrast techniques, pre-procedural optimization protocols, and post-procedural monitoring strategies.

#### 4. CKD-specific risk prediction models

Development and validation of risk calculators incorporating renal function parameters, frailty measures, and procedural variables.

#### 5. Health economic analyses

Cost-effectiveness studies comparing TAVI vs. SAVR in CKD populations, including quality-of-life measures and healthcare resource utilization.

### Conclusions

This comprehensive systematic review demonstrates that chronic kidney disease is associated with higher procedural and intermediate-term risks following TAVI, particularly for mortality and acute kidney injury. However, the majority of CKD patients experience stable or improved renal function post-procedure, supporting the appropriateness of TAVI in carefully selected patients with renal impairment.

The evidence supports a multidisciplinary approach to CKD patients being considered for TAVI, with emphasis on pre-procedural optimization, renal-protective procedural strategies, and structured post-procedural monitoring. Future research should focus on standardizing outcome definitions, investigating long-term valve durability, and developing CKD-specific risk prediction models to optimize patient selection and outcomes.

## Data Availability

All data produced in the present study are available upon reasonable request to the authors

## Acknowledgments

The authors acknowledge the contributions of all researchers whose work was included in this systematic review.

## Data Availability Statement

All data supporting the conclusions of this systematic review are available in the published literature cited herein.

